# Genetics of the Leading Causes of Death in Human Aging

**DOI:** 10.64898/2026.05.04.26352398

**Authors:** Alexandria Martignoni, Wyen C Cai, Victoria Calderon, Crystal C Aguinaldo, Kenneth Park, Shin Murakami

**Affiliations:** Department of Foundational Biomedical Sciences, Touro Interdisciplinary Institute for Healthy Aging, College of Osteopathic Medicine, Touro University California, Vallejo, CA; Biology Department, Napa Valley College, Napa, CA, United States

**Keywords:** Aging, Leading causes of death, Genetic contributions, Cardiovascular disease, Stroke, Cancer, Diabetes, Immune and inflammatory diseases, Metabolic diseases, Stress resistance

## Abstract

The relationship between age-related genetic factors and health conditions has become a pivotal focus in aging research, particularly as the World Health Organization (WHO) delineates the leading global causes of mortality. However, the direct impact of age-related genes on the leading cause of death remains poorly understood. To investigate this gene-aging relationship, we analyzed protein-protein interactions using gene set enrichment analysis (GSEA) of a set of 307 age-related genes previously curated. The results indicated significant associations with 113 diverse disease categories, while adhering to a stringent false discovery rate (FDR) threshold of less than 1 x 10^-5^. Due to the difficulties in aligning the disease categories with WHO’s leading causes of death, we reclassified the WHO categories using the more precise nomenclature specified in the 11^th^ Revision of the International Classification of Diseases (ICD-11). The age-related genes account for the leading causes of death, with the exceptions being two infectious communicable diseases, tuberculosis and COVID-19. They impact the cardiovascular system, brain, lungs, and the whole body, while this study could not identify death by aging, which is not a well-defined medical cause of death. Furthermore, we identified a set of 15 recurring genes shared among multiple diseases, including TNF, AKT1, IL6, CDK2A, APOE, and TP53. This gene set was enriched for several disease categories, including cancer, inflammatory diseases, metabolic disorders, and neurodegenerative diseases. Additionally, it shows significant enrichment in various biological categories, with the regulation of nitric oxide activity being the most prominent; other enriched categories include the regulation of microRNA, lipid and carbohydrate metabolism, smooth muscle cell proliferation, insulin signaling, and phosphatidylinositol-3 kinase (PI3K) signaling. The findings suggest that the recurring genes act as pleiotropic hubs, influencing multiple leading causes of death, while other genes are more specific to each disease category.

## 1. Introduction

The global mortality trends related to the leading causes of death are crucial for understanding and advancing public health initiatives and mitigating disease burden. The World Health Organization (WHO) reported that, in 2021, the foremost contributors to global mortality included ischemic heart disease, COVID-19, cerebrovascular accidents (strokes), chronic obstructive pulmonary disease (COPD), lower respiratory infections, lung malignancies, dementia, diabetes mellitus, renal disease, and tuberculosis [1,2]. In the United States, the leading causes mirror some of the global trends but also highlight unique factors, such as ischemic heart disease, dementia, COPD, substance use disorders, cerebrovascular accidents, tracheobronchial cancers, renal pathologies, hypertensive heart disease, and self-inflicted injuries [3]. The WHO aggregates mortality data on these causes by systematically monitoring annual death rates, classifying them into three distinct categories: communicable diseases, noncommunicable diseases, and injuries, enabling health authorities to identify priority areas for intervention and resource allocation [1,2].

In previous studies, we showed the genetic factors associated with Alzheimer’s disease, which is a significant cause of mortality in individuals over 65 years old [4,5]. In this study, we aim to broaden our study to include the top 10 leading causes of death as defined by the World Health Organization (WHO). We investigated both communicable and non-communicable diseases, using the validated genes associated with aging (referred to as age-related genes) in humans. Communicable diseases are those transmitted from person to person via various modes due to pathogens such as viruses or bacteria. Notable examples include COVID-19, respiratory infections, and tuberculosis [5,6]. Lower respiratory tract infections (LRTIs) rank among the WHO’s top ten global causes of mortality and are classified under section 12 of the International Classification of Diseases, 11th Revision (ICD-11) [7], which addresses Diseases of the Respiratory System. Tuberculosis (TB), a significant global health concern, is classified among the top causes of death by the WHO and falls under section 1 in the ICD-11 [7], which covers Certain Infectious or Parasitic Diseases. The etiological agent, Mycobacterium tuberculosis, primarily targets the pulmonary system [8]. COVID-19 was identified as the second leading cause of death worldwide in 2021 and is categorized in section 25 of the ICD-11, designated for Codes for Special Purposes [7]. In contrast, non-communicable diseases (NCDs), often referred to as chronic diseases, typically manifest later in life. These conditions include ischemic heart disease, stroke, chronic obstructive pulmonary disease, lung cancers, diabetes mellitus, and chronic kidney disease. NCDs are responsible for approximately 74% of global mortality, with cardiovascular diseases representing the most lethal category, accounting for about 44% of these deaths [9]. We then identified health conditions that are directly associated with age-related genes using GSGA.

## 2. Methods

### 2.1. Data Collection and Processing, Gene Set Gene Enrichment analysis (GSEA), Over-representation Analysis (ORA), and Statistical Analysis

A validated list of 307 age-related (AR) genes described in the previous study [10] was used in this analysis. This gene list was then imported into the STRING-DB version 12.0 [11] (https://string-db.org/; Last accessed on 10/4/2025) to perform GSEA as described previously [5]. Statistical analysis was performed using the disease enrichment analysis tool from GSEA, which incorporates an integrated confidence scoring system to assess the strength of gene-disease associations. The final dataset comprised AR genes, their related diseases, and categorized disease information. This dataset was analyzed to uncover patterns of disease associated with AR genetic pathways. To refine the dataset, a false discovery rate (FDR) threshold of <= 1 x 10^-5^ was applied to determine the significance of these associations. Diseases with an FDR below this criterion were retained for subsequent analyses, ensuring the focus on statistically robust links to AR genes. For result outputs, we used network presentation, functional enrichments in your network with the enrichment display. We subsequently aligned disease categories that are directly correlated with age-related genes using Gene Set Gene Enrichment analysis (GSGA).

To further delineate the functional composition of our age-related gene dataset, we conducted over-representation analysis (ORA) of the gene set across gene ontology (GO) domains, including Biological Process, Cellular Component, and Molecular Function categories. ORA was conducted using WebGestalt (WEB-based Gene Set Analysis Toolkit, Last accessed 5/3/2026) [12] with Homo sapiens selected as the organism and official gene symbols as identifiers. ORA was performed with disease category significance level set to an FDR threshold of ≤ 0.05. Of the original validated dataset of 307 age-related genes, ORA included 302 genes with available GO annotations. To evaluate disease associations, disease ORA was performed in WebGestalt using DisGeNET disease annotations with all protein-coding genes as the reference set. Only disease categories containing a minimum of three genes were included, and redundancy among disease terms was reduced using weighted set cover.

### 2.2 Disease Categorization

Diseases were categorized as either communicable or noncommunicable to highlight their prevalence. This framework was subsequently grouped to correlate with the eight primary categories derived from the WHO’s leading global causes of mortality [1]. They were further classified based on their pathophysiological mechanisms and the organ systems involved, aligning with the International Classification of Diseases, 11th Revision (ICD-11) [7]. Gene-disease associations identified with a false discovery rate (FDR) of less than 1 x 10^-5^ were distinguished under the following global causes of death: (1) Lower Respiratory Infections, (2) Cancers of the Trachea, Bronchus, and Lung, (3) Chronic Obstructive Pulmonary Disease, (4) Ischemic Heart Disease, (5) Stroke, (6) Alzheimer’s Disease and other Dementias, (7) Diabetes Mellitus, and (8) Kidney Disease.

### 2.3 Condensed Gene Set Analysis

Further analysis was conducted by importing the fifteen most recurring genes as a condensed gene list into STRING-DB as described above. Functional enrichment and clustering analyses were performed using default STRING parameters, and disease annotation results were obtained using STRING disease databases. The MCL (Markov Clustering) embedded in the STRING-DB was used to create an interaction network [11].

## 3. Results

Using a validated dataset of 307 age-related genes, we conducted GSEA and identified 113 disease categories linked to these genes (refer to Supplementary Table 1). Of the 113 diseases identified by GSEA using an FDR of < 1 x 10^-5^, a total of 25 diseases (referred to as subcategories) were associated with the WHO’s leading causes of death. However, these disease categories included a wide range of disease dimensions and ontological terminologies, for example, broadly ranging from cellular, tissue, and organ to systemic levels, and thus, it was not straightforward to align with the WHO’s leading causes of death. Consequently, we undertook an analysis to reclassify the WHO categories using a more precise nomenclature as specified in the International Classification of Diseases (ICD-11).

### 3.1 Classification of WHO disease categories

A key challenge in this study was aligning the World Health Organization’s (WHO) terminology with genetic ontological categories. The WHO’s leading causes of death use medical terms within an epidemiological framework, with the underlying causes coded using the International Classification of Diseases, 11th revision (ICD-11). To correlate the categories of genetic diseases with the leading causes of death, we classified the ICD-11 categories. Among the WHO’s leading causes of death, we identified eight major categories. These categories were linked to the top eight ICD-11 classifications for leading causes of death, followed by further research to ensure proper categorization (Supplementary Table 1).

Figure 1 summarizes the disease categories. The ICD-11 categorizes diseases into communicable and noncommunicable groups based on their prevalence and association. Within the category of communicable diseases, Lower Respiratory Tract Infections (LRTI) encompass significant health concerns, such as tuberculosis (TB) and COVID-19, both of which are known to cause lower respiratory tract infections. Although the World Health Organization (WHO) ranks these diseases among the top global causes of death, they have been included under LRTI due to their manifestation in the respiratory system. Chronic Obstructive Pulmonary Disease (COPD) is corroborated by affiliations with lower respiratory tract diseases, emphasizing its impact on lung health. Notably, respiratory tract and lower respiratory tract diseases (RT and LRT) share symptoms among three categories: one communicable disease category (lower respiratory infections), two non-communicable disease categories (chronic obstructive pulmonary disease, and trachea, bronchus, and lung cancer) (Figure 1).

**Figure 1.**
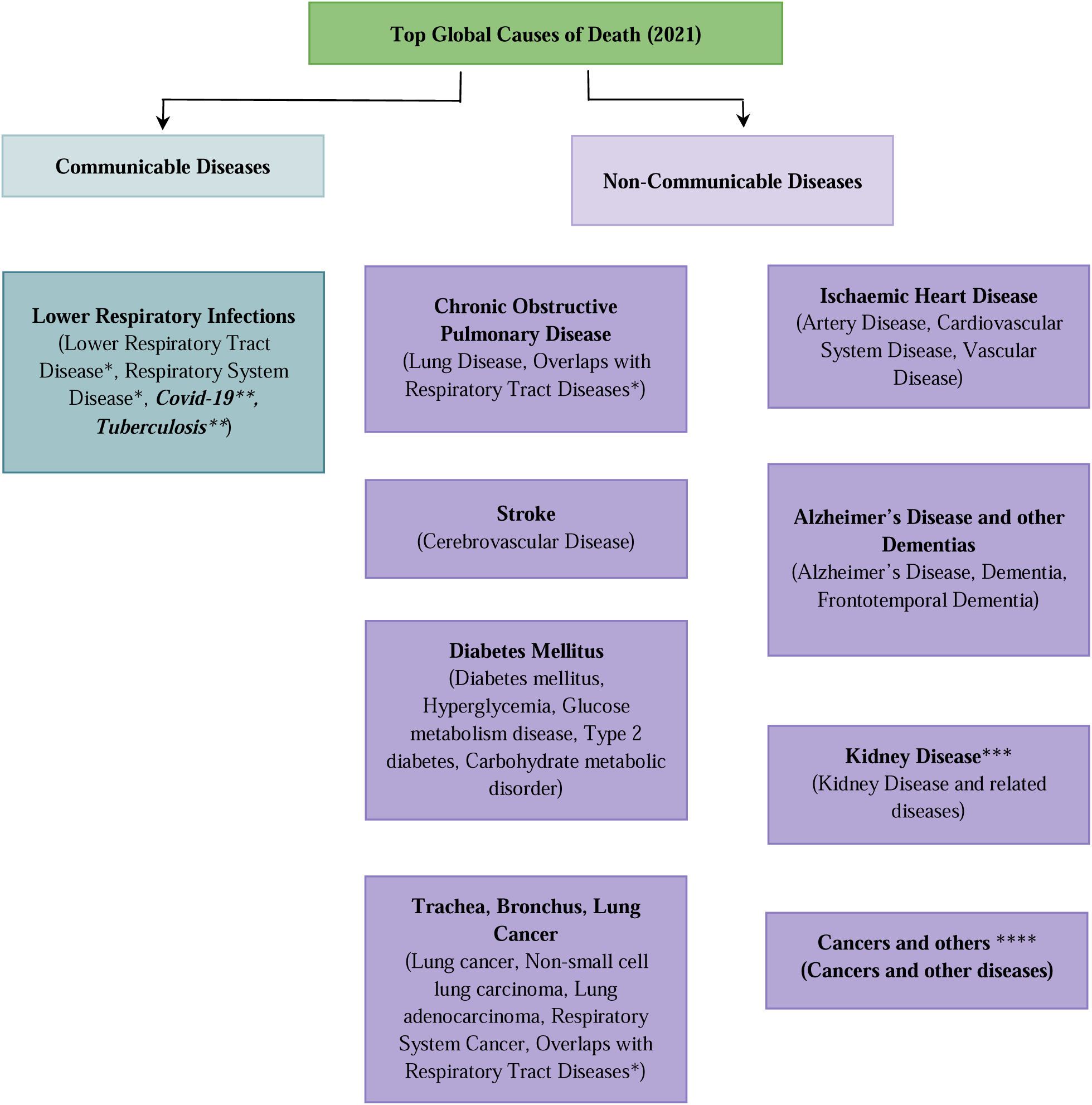
The WHO’s 2021 list of top global causes of death aligns with the disease categories defined by this study using GSEA. Communicable diseases are indicated by blue-green; Non-communicable diseases are indicated by purple. Note that cancer has been identified by this study, which was not included in the WHO’s top 10 leading causes of death. * Due to overlapping symptoms, respiratory tract diseases are classified into three categories: lower respiratory infections, chronic obstructive pulmonary disease, and trachea, bronchus, and lung cancer. ** Two infectious diseases in the communicable disease category, COVID-19 and Tuberculosis, were not identified by this study. *** The category of kidney disease is under the ICD-11 genitourinary system, which can influence kidney functions. **** The category of cancers and other diseases includes those that were not specific to the other categories but could influence them.

Noncommunicable diseases include a range of conditions with varying prevalence and genetic associations. For example, Chronic Obstructive Pulmonary Disease has been linked to lower respiratory tract diseases, while cancers affecting the trachea, bronchus, and lungs have demonstrated substantial gene associations as described below. The data reveal multiple forms of lung cancer—including non-small cell lung carcinoma and lung adenocarcinoma—with significant FDR values, confirming the complexity and severity of respiratory system cancers. Cancers related to the category of trachea, bronchus, and lung as well as cancers in general, were classified in different categories of ICD-11 and therefore we included them in two separate categories, one category was leading causes of death, trachea, bronchus, and lung cancers and another categories was cancers and others (Figure 1). Ischemic Heart Disease, a leading cause of mortality, is associated with several vascular diseases, highlighting the relationship between arterial health and cardiovascular risks.

Stroke and dementia-related diseases further illustrate the diversity of noncommunicable diseases. Stroke ranks as the second leading cause of death globally, categorized under diseases of the nervous system, and possesses distinct gene associations pertaining to cerebrovascular health. Alzheimer’s Disease and other dementias show varied prevalence and genetic factors, underscoring the importance of understanding cognitive health conditions. Diabetes mellitus, also categorized under noncommunicable diseases, demonstrates strong gene correlations as well and is critical in public health discussions surrounding metabolic disorders. Overall, these classifications and their related data underscore the complex relationships between various diseases and emphasize the need for continued research and awareness in managing both communicable and noncommunicable diseases.

### 3.2 Detailed results of ICD-11 diseases aligned with GSEA categories

Table 1 summarizes the details of ICD-11 diseases aligned to the GSEA results. As shown in Figure 1, the ICD-11 diseases were divided into either communicable or noncommunicable, to highlight their prevalence.

**Table 1.**
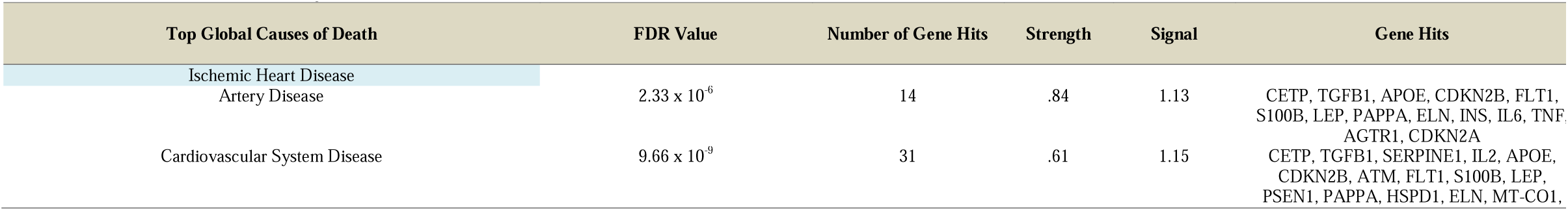
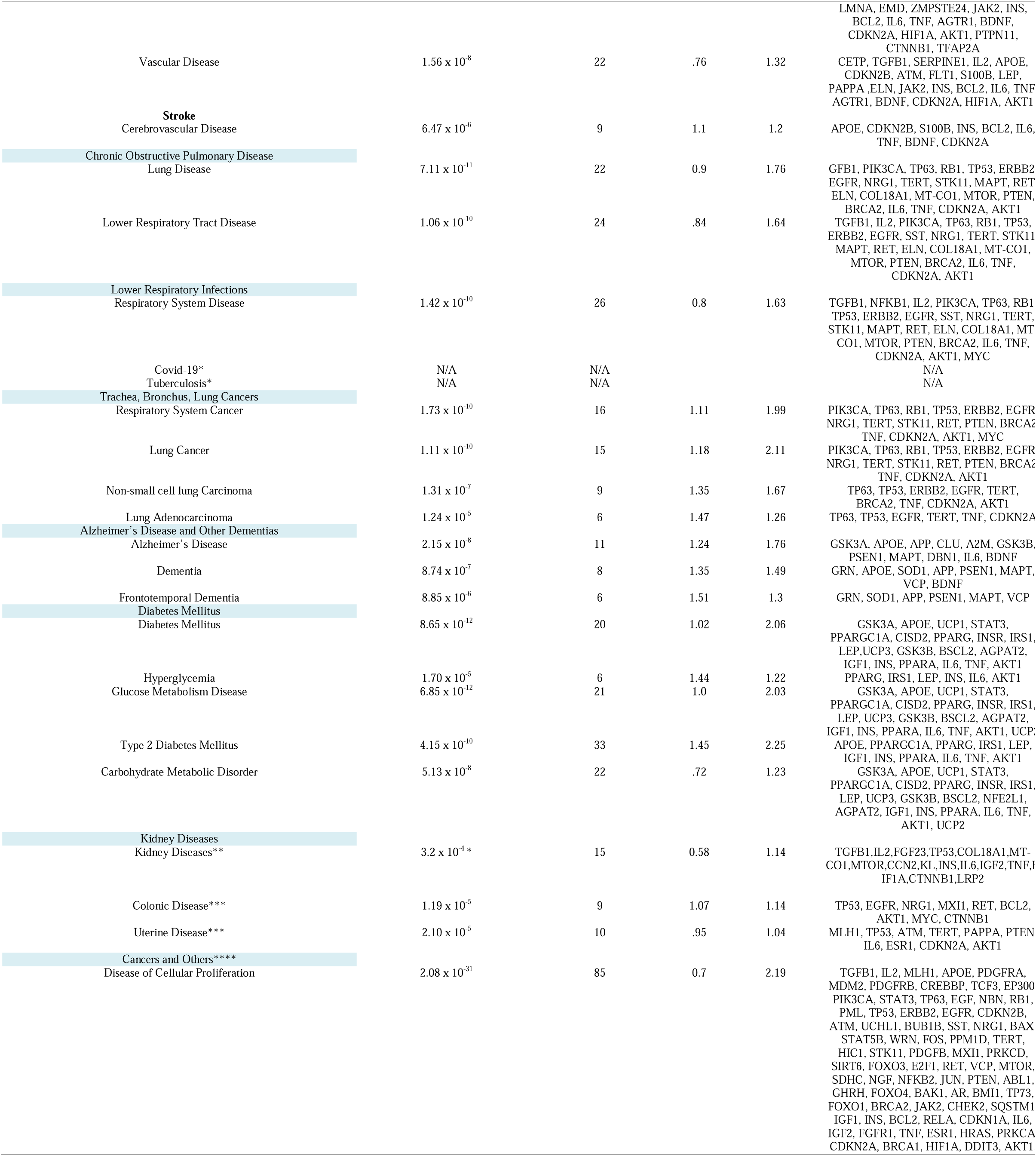
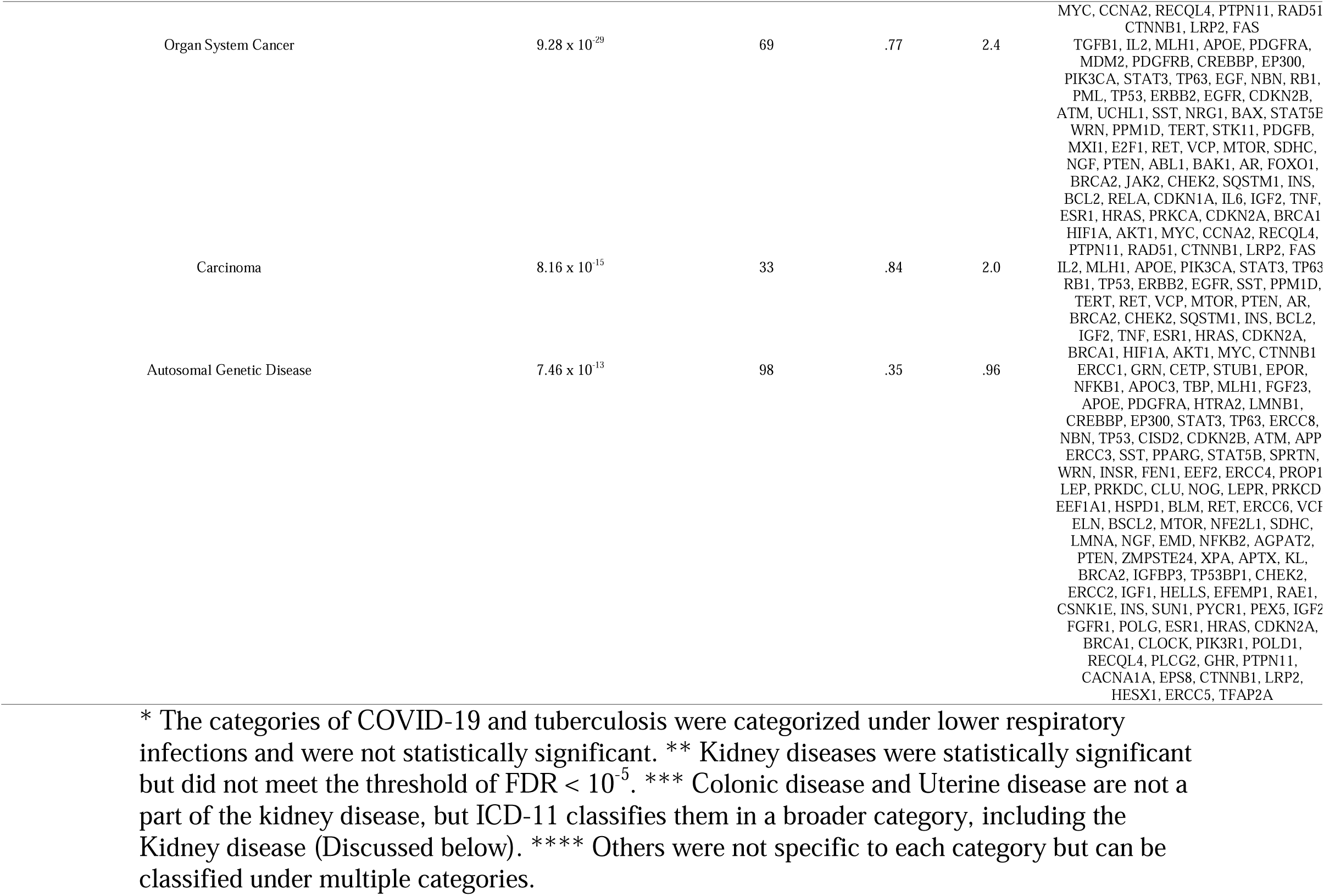
Top global causes of death with diseases, FDR values, and gene hits. The disease categories were identified by GSEA.

#### 3.2.1 Communicable Diseases

##### Lower Respiratory Tract Infections

Respiratory system disease had an FDR of 1.42 x 10^-10^ with 27 gene hits. TB does not have its own category under GSEA due to a lack of a strong association with other GSEA diseases and thus does not have an FDR. Due to the relatively transient nature of the disease, COVID-19, which in part may be involved here, does not have its own category under GSEA and thus lacks an FDR.

#### 3.2.2 Noncommunicable Diseases

Seven categories are classified into this group as follows:

##### Chronic Obstructive Pulmonary Disease

This category includes lower respiratory tract diseases (LRTD), which had an FDR of 1.06 x 10^-10^ with 25 gene hits. A general category, lung disease, was statistically significant (FDR of 7.11 x 10^-11^; 22 gene hits) that includes Chronic Obstructive Pulmonary Disease.

##### Trachea, Bronchus, and Lung Cancers

RS cancer in this category had an FDR of 1.73 x 10^-10^ with 16 gene hits. Lung cancer had an FDR of 1.11 x 10^-10^ with 15 gene hits. Non-small cell lung carcinoma (NSCLC) had an FDR of 1.31 x 10^-7^ with 9 gene hits. Lung adenocarcinoma had an FDR of 1.24 x 10^-5^ with 6 gene hits. The ICD-11 categorizes lung adenocarcinoma under section 2. Lung cancer is the most common form of adenocarcinoma, further supporting its alignment with TBL cancers [12].

##### Ischemic Heart Disease

Artery disease had an FDR rate of 2.33 x 10^-6^ with 14 gene hits. The ICD-11 puts artery disease under section 11. Cardiovascular system disease had an FDR of 9.66 x 10^-9^ with 29 gene hits. Vascular disease had an FDR rate of 1.56 x 10^-8^ with 22 gene hits.

##### Stroke

The second leading cause of death globally, according to the WHO, is stroke and is in the ICD-11 under section 8 - Diseases of the Nervous System. Cerebrovascular disease had an FDR of 6.47 x 10^-6^ with 9 gene hits, and the ICD-11 categorizes the disease under category 8.

##### Alzheimer’s Disease and Other Dementias

Alzheimer’s disease had an FDR of 2.15 x 10^-8^ with 11 gene hits. Dementia FDR rate was 8.74 x 10^-7^ with 8 hits. Frontotemporal dementia FDR rate was 8.85 x 10^-6^ with 6 gene hits.

##### Diabetes Mellitus

DM has an FDR of 8.65 x 10^-12^ with 20 gene hits and falls under ICD-11 section 5. Hyperglycemia FDR was 1.70 x 10^-12^ with 6 gene hits. Glucose metabolism disease FDR was 6.85 x 10^-12^ with 21 gene hits. Type II diabetes (T2D) FDR was 4.15 x 10^-10^ with 11 gene hits and can be found under section 5. Carbohydrate metabolism disorder had an FDR of 5.13 x 10^-8^ with 22 gene hits.

##### Kidney Disease, Colonic Disease, and Uterine Disease

ICD-11 Section 16 classifies conditions related to the genitourinary system, which includes diseases of the kidneys and reproductive organs [7]. It is important to note that colonic and uterine diseases can affect kidney function, which is why they are included in this category.

While the categories of kidney diseases showed statistically significant associations, they did not meet our predetermined threshold for the false discovery rate (FDR) of less than 10^-5. Specifically, kidney diseases had an FDR of 3.2 x 10^-4, leading us to exclude the gene set enrichment analysis (GSEA) category for kidney disease from further analysis.

On the other hand, colonic diseases, such as colon cancer, demonstrated a significant association with an FDR of 1.19 x 10^-5^ and included 9 instances. Uterine diseases, which were also classified under the genitourinary system (as mentioned concerning kidney disease), had an FDR of 2.10 x 10^-5 and comprised 10 instances.

##### Cancers and other diseases

The vast majority of diseases in this category were not straightforward to assign to a specific category and overlapped with multiple diseases, particularly with the category of cancer. The category of the disease of cellular proliferation, including various types of cancers, results from abnormal cell division. DCP had an FDR of 2.08 x 10^-31^ with 29 gene hits. Organ system cancer had an FDR of 9.28 x 10^-29^ with 29 gene hits. Carcinoma had an FDR of 8.16 x 10^-15^ with 31 gene hits.

We included autosomal genetic disease in this category, which largely overlaps with cancers and leading causes of death, with an FDR of 7.46 x 10^-13^. This category ranges broadly with 98 gene hits enriched with cancers or diseases with cancer outcomes (prostate cancer, reproductive organ cancers, and Xeroderma pigmentosum), accelerated aging syndromes (Cockayne syndrome and progeria) (FDR < 1.51 x 10^-9^), among others. The biological mechanisms mostly cover responses and resistance to multiple forms of stress and stimuli (Cellular response to stimulus, FDR = 1.03 x 10^-25^; DNA damage response and repair, FDR < 1.12 x 10^-19^; Response to radiation, FDR = 9.45×10^-21^; Regulation of cell death, FDR = 6.07 x 10^-18^) and metabolic process (nitrogen compound metabolic process, FDR = 17.79 x 10^-23^; Cellular metabolic process, FDR = 1.08 x 10^-18^).

### 3.3 Identification and GSEA of Highly Recurrent Genes Across Diseases

Fifteen genes were identified as recurring across multiple disease categories (Figure 2). These genes were selected as a condensed gene list for subsequent network-based and enrichment analyses. Table 2 summarizes the results. Enrichment using the condensed gene set showed a shift toward the following diseases (categories, FDR): Cancer (carcinoma, 1.95 x 10^-16^; disease of cellular proliferation, 1.10 x 10^-14^; organ system cancer, 3.37 x 10^-15^); lung, breast ad large intestine, and other cancers, <7.13 x 10^-6^), inflammatory disease (Colonic Disease, 1.03 x 10-5); diabetes, metabolic and gastrointestinal diseases (type 2 diabetes mellitus, 6.64 x 10^-12^, carbohydrate metabolic disorder, 1.09 x 10^-8^, pancreas disease, 4.59 x 10^-18^, among others (Supplementary Table 2). They impact a wide range of organ systems, including the lung, breast, gastrointestine (pancreas, stomach, and large intestine), endocrine, vascular, among others (Supplementary Table 2).

**Figure 2.**
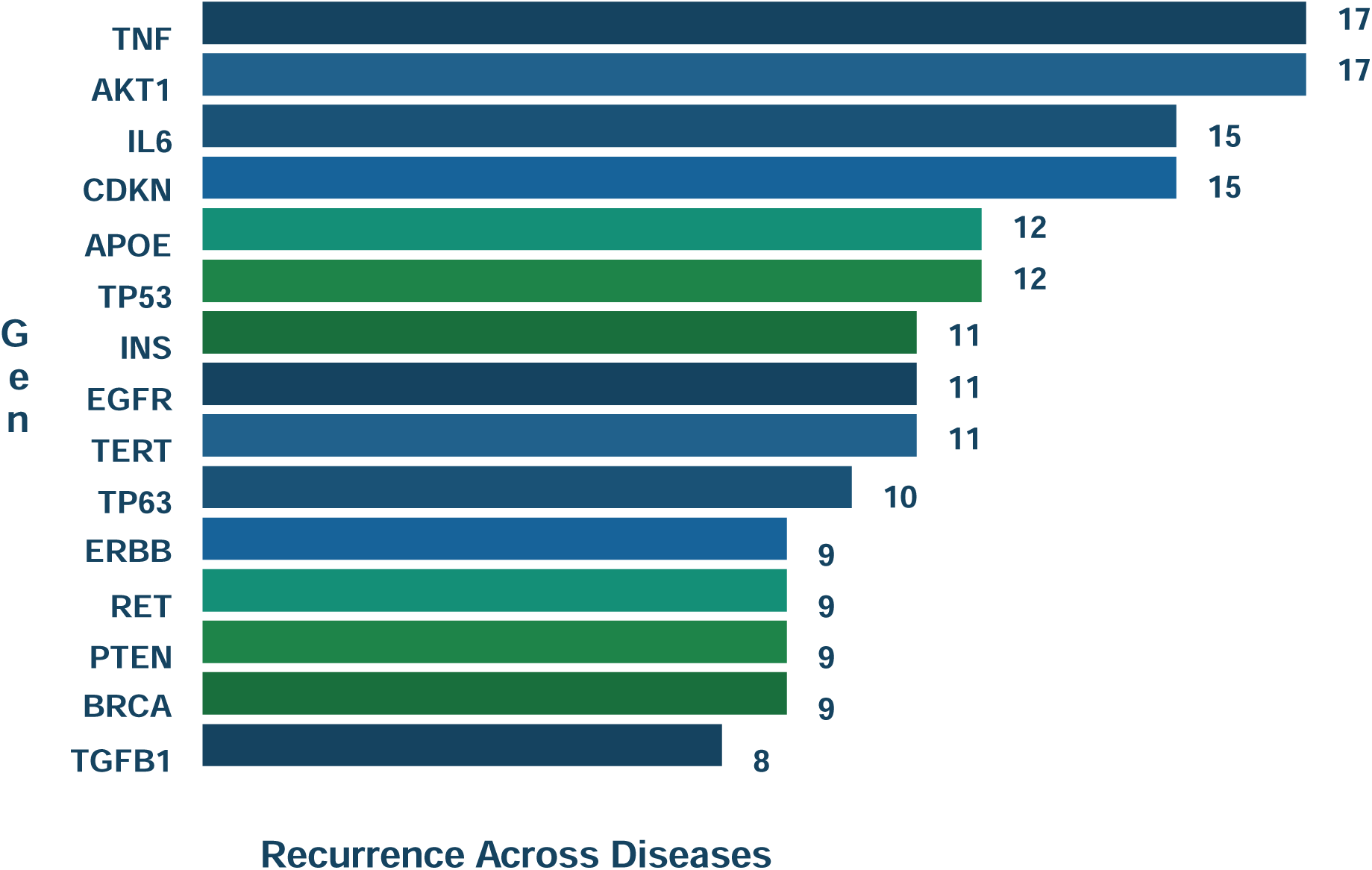
Top 15 genes recurring across multiple disease categories.

Furthermore, the biological network-based functional clustering highlights the regulation of nitric-oxide activities (FDR = 2.73 x 10^-11^) as the primary category (Figure 3), which is involved in the inflammatory and vascular regulatory processes. Other categories include regulation of microRNA (FDR < 1.47 x 10^-7^), lipid and carbohydrate metabolism (FDR < 9.84 x 10^-8^), regulation of smooth muscle cell proliferation (FDR = 2.35 x 10^-9^), and regulation of phosphatidylinositol-3 kinase (PI3K) signaling (FDR = 3.05 x 10^-8^).

**Figure 3.**
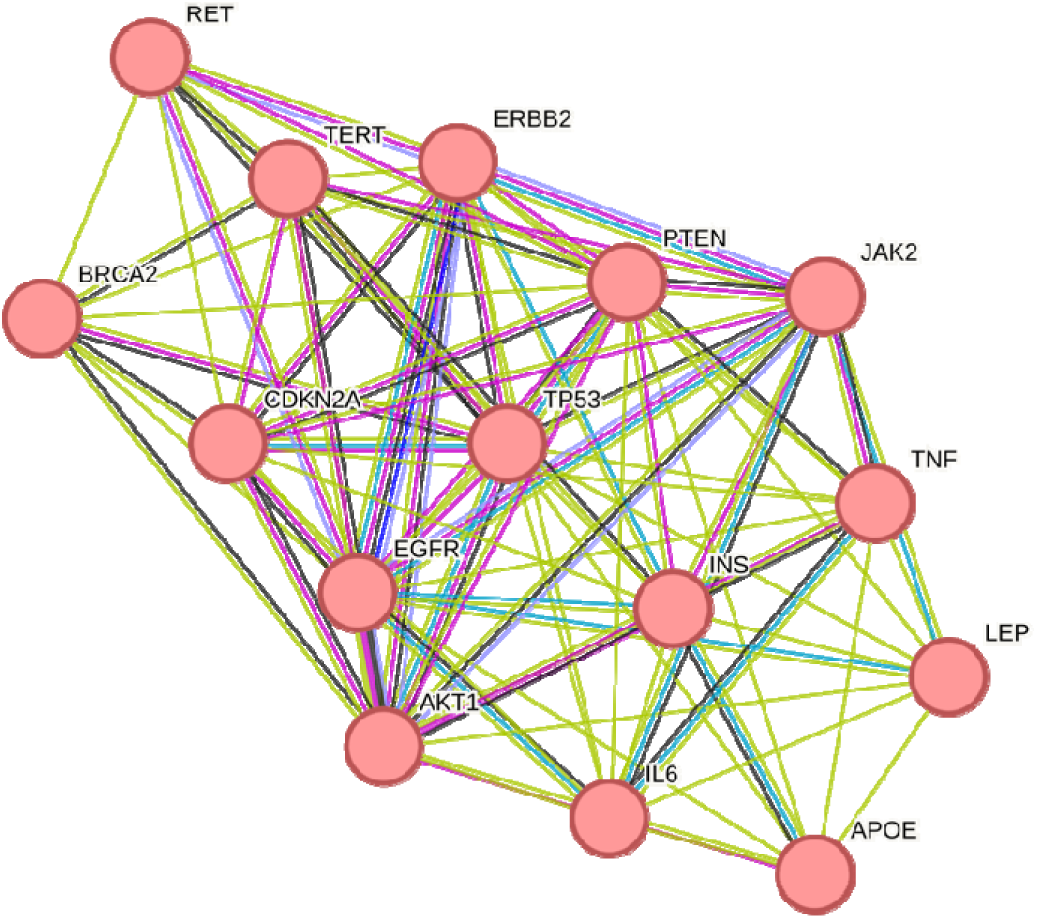
The major cluster among the recurring 15 age-related genes. The MCL (Markov Clustering) analysis highlighted the regulation of nitric-oxide synthase activity as a major group.

### 3.4 Disease Over-Representation Analysis Using WebGestalt

Disease ORA using WebGestalt (DisGeNET) revealed significant enrichment of neoplastic, cardiovascular, metabolic, and neurodegenerative disease categories (Figure 4). Cancer-related terms such as carcinoma dominated the enrichment profile, reflecting overrepresentation of core oncogenic and tumor suppressor genes (e.g., TP53, AKT1, PTEN, BRCA2, EGFR).

**Figure 4.**
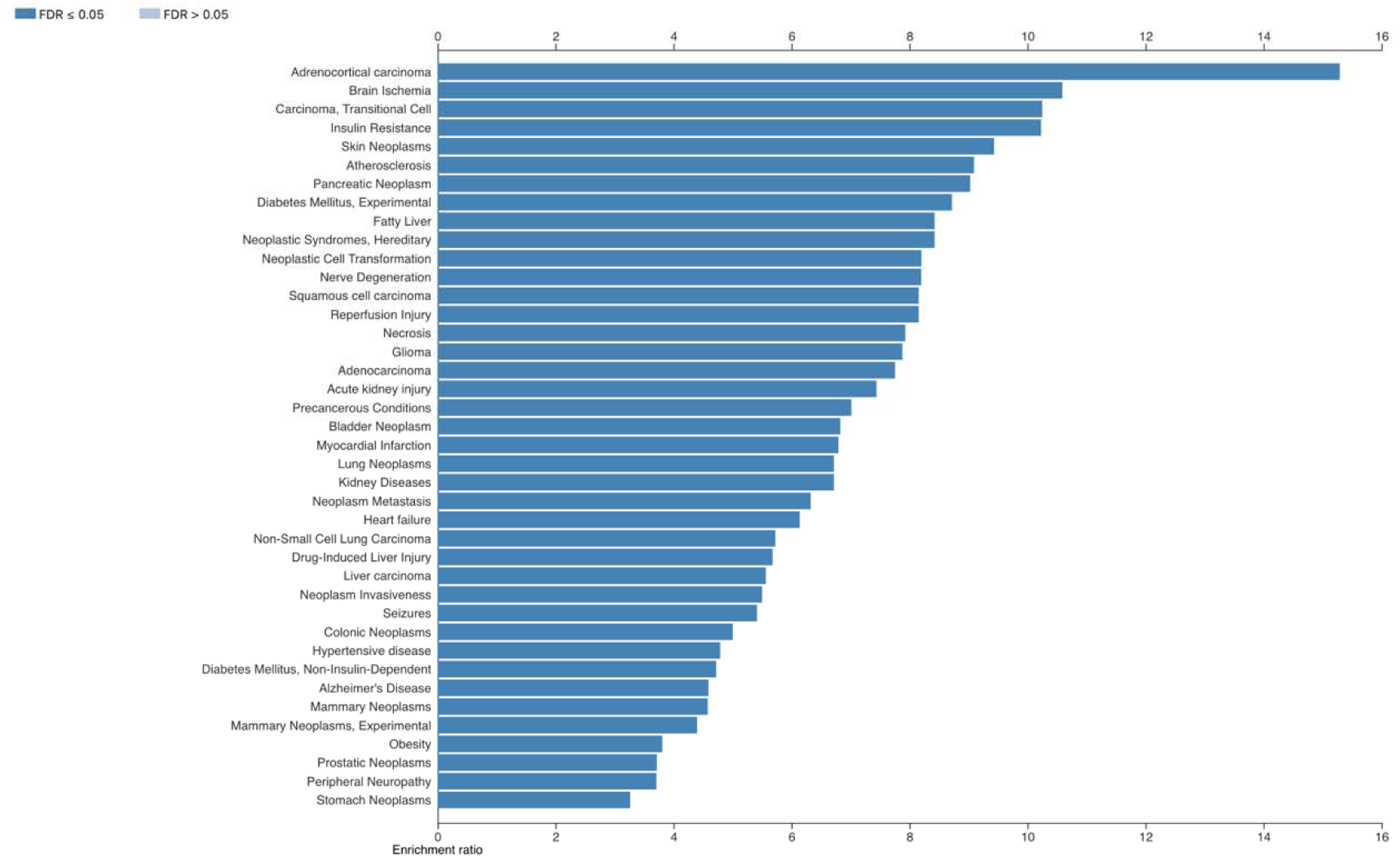
Disease over-representation analysis (ORA) of recurrent genes using WebGestalt (DisGeNET). Bars represent enrichment ratio. Only disease categories with FDR ≤ 0.05 are shown.

The functional composition of age-related genes is as follows. Gene ontology (GO) classification demonstrated that age-related genes are predominantly associated with biological regulation, metabolic processes, and cellular responses to stimuli (Figure 5). Cellular component analysis indicated predominant localization to the nucleus, cytoplasm, and membrane-enclosed compartments, while molecular function categories were dominated by protein binding and enzymatic activities.

**Figure 5.**
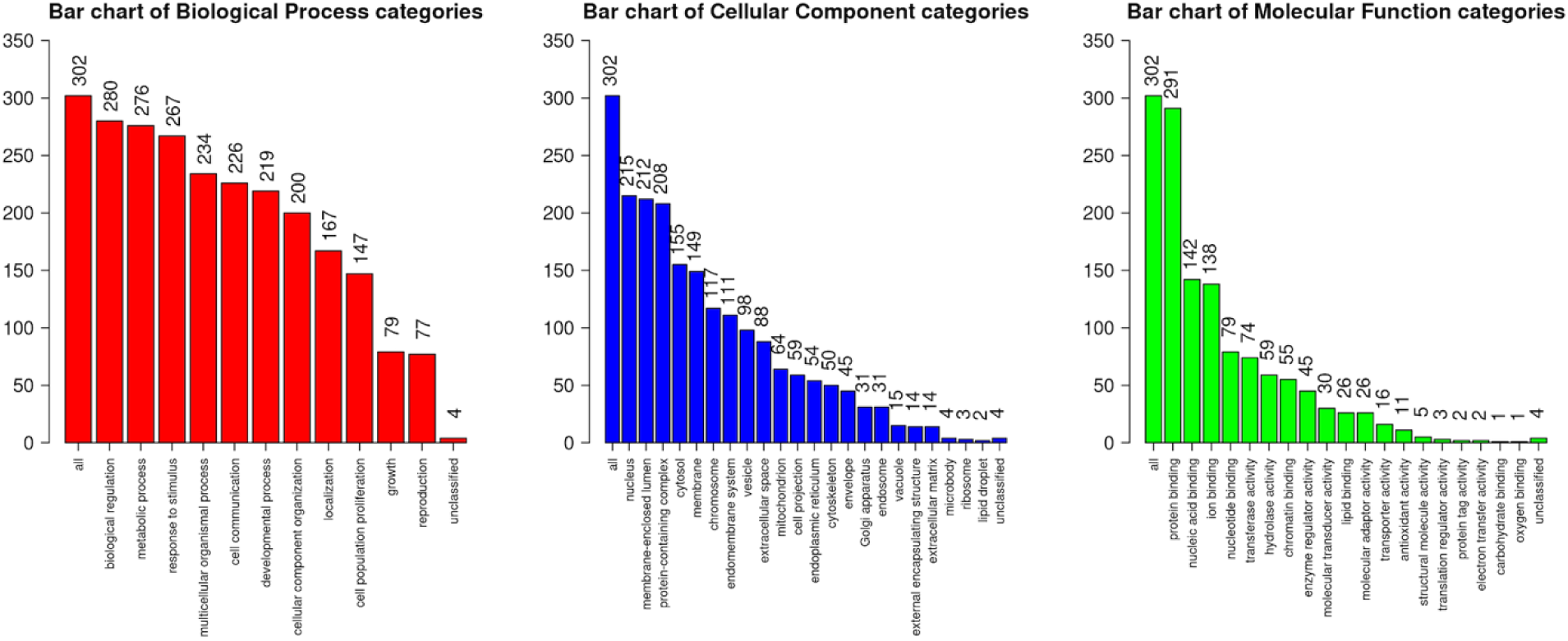
Over-representation analysis (ORA) of the gene distributions, using Gene Ontology (GO) biological process (red), cellular component (blue), and molecular function (green) categories, as shown in the figure, with numbers indicating the genes annotated to each category.

## 4. Discussion

### 4.1. Overview of the disease categories extracted from age-related genes

This study investigates the relationship between age-associated genetic factors and major global mortality contributors, employing GSEA for a categorization schema that visually delineates varying correlation strengths. The results identified 113 diverse disease categories, accounting for 8 out of 10 categories plus 1 cancer category not included in the WHO categories; the categories not covered by the genes are two infectious diseases, tuberculosis and COVID-19. The results suggest that the age-related genes influence all the non-communicable disease (NCD) categories among the leading causes of death.

We further identified 15 age-related genes that are common across multiple diseases. The genes most frequently identified in the GSEA disease categories, TNF and AKT1, followed by IL6, CDK2A, APOE, and TP53, among others. While AKT1 has shown no significant correlation with longevity in Danish and German cohorts of non-centenarians and centenarians [13], the model-system studies in mice, fruit flies, and soil nematodes indicate a direct involvement of AKT in life-extending mechanisms [14]. For example, a murine study suggests that haploinsufficiency of AKT1 may enhance lifespan by reducing energy expenditure and oxidative stress [15].

Importantly, the functional clustering of the 15 recurring genes highlights regulation of nitric oxide activity as the primary category, followed by regulation of microRNA (miRNA), lipid and carbohydrate metabolism, smooth muscle cell proliferation, insulin signaling, and phosphatidylinositol-3 kinase (PI3K) signaling. These findings suggest that there are shared biological mechanisms among the leading causes of death. Firstly, Nitric oxide (NO) production decreases by half from age 20 to age 40 [16,17]. However, the expression of endothelial NOS (eNOS), which indicates NO levels in the vascular endothelium, has been a subject of debate [18]. The age-related decline in nitric oxide production can lead to arterial stiffening, reduced cognitive performance, and an increased risk of cardiovascular disease [16,18,19].

The NO production declines at the age of 40 years aligns with common midlife issues, particularly metabolic problems that may arise around the ages of 44 and 60, despite its relatively small sample size [20]. A systematic review identifies that the onset of the risks for type 2 diabetes peaks at 55-60 years old [21], while Alzheimer’s disease becomes prevalent after 65 years old [22,23]. In fact, this study identified lipid and carbohydrate metabolism as significant biological categories. Furthermore, the insulin signaling pathway involves key components such as phosphatidylinositol-3 kinase (PI3K), AKT, and the FOXO transcription factor. Together, these elements form an insulin/IGF-1 signal transduction pathway that plays a crucial role in determining lifespan and responding to various forms of stress in model organisms, including mice, fruit flies, and soil nematodes [14,24].

Secondly, MicroRNA is a type of non-coding small RNA that is believed to play a role in aging [25–27]. A well-known example of its function is the suppression of P53 through miR-33, miR-125b, and miR-504, while miR-34 and miR-125a/b are known to activate P53 [27]. In model system studies involving soil nematodes and fruit flies, the orthologous genes of miR-125a/b have been shown to shorten lifespans, whereas the orthologous genes of miR-183 and miR-34a/b/c extend lifespan in nematodes but shorten it in fruit flies [24,25]. There is inconsistency regarding the effects of these genes among different species, and as a result, the precise regulatory mechanisms of microRNA require further investigation.

The recurring genes also highlight shared disease categories, including cancer, inflammatory diseases, metabolic disorders, and neurodegenerative diseases, particularly neoplasms and inflammatory diseases. Neoplasms are the leading cause of death in the United States [3], even though cancer does not appear among the top ten WHO’s leading causes of death. Our research revealed that a wide variety of malignancies—specifically, tumor, blood, and lymphoid (TBL) neoplasms and their subtypes—show a strong correlation with age-related genetic variants. This disease category can affect a wide variety of tissues and organs and therefore, it was placed in a separate category (Figure 1).

There is a well-documented relationship between compromised immune function and an increased risk of cancer [28]. This decline in the immune system’s ability to combat cancer as individuals age is referred to as immune senescence [29]. Furthermore, the immune system’s functionality is closely tied to disease susceptibility. As people age, the immune system tends to become more pro-inflammatory, while its effectiveness diminishes. This decline is likely due to reduced responsiveness from B and T lymphocytes [30].

#### 4.1.1. Specific disease categories for the leading causes of death

##### 4.1.1.1 Lower Respiratory Tract Infections (LRTI)

Lower Respiratory Tract Infections (LRTIs) represent a group of infectious diseases primarily affecting the lower respiratory tract, distinct from upper respiratory conditions. LRTIs can stem from various etiologies, including infectious agents, genetic predispositions, and pathological processes that impair pulmonary function (National Institute for Health and Clinical Excellence, 2008). Our results were consistent with genetic contributions to LRTIs, which were classified into the three categories of the leading causes of death due to shared symptoms (Figure 1). Although our GSEA did not specifically identify categories of tuberculosis and COVID-19, existing research has illuminated several pivotal immune mediators implicated in TB pathogenesis. Key factors include Interleukin-6 (IL-6), Tumor Necrosis Factor (TNF), Interleukin-2 (IL-2), NFKB1, and Transforming Growth Factor Beta 1 (TGFB1), all integral to the immune dynamics of tuberculosis and related respiratory conditions. IL-6 is notably elevated in TB patients [31], whereas diminished TNF levels correlate with accelerated disease progression [32]. Furthermore, IL-2 conspicuously increases during active TB infections, underscoring its critical role in orchestrating immune responses [33,34].

##### 4.1.1.2 Chronic Obstructive Disease (COPD)

Chronic obstructive pulmonary disease (COPD) is a leading cause of mortality worldwide, according to the World Health Organization (WHO). This progressive disorder is characterized by airway obstruction, lung tissue degradation, and pulmonary vascular changes, leading to airflow limitation. COPD falls under Section 12 of the WHO’s International Classification of Diseases (ICD-11) [3]. Thus, we used this disease category. The section also includes lower respiratory tract diseases (LRTD), which encompass prevalent conditions such as asthma [35]. Age is a significant risk factor due to the disease’s gradual progression [36]. COPD and asthma are both associated with airway narrowing and expiratory dyspnea [37]. Our study identified key genes linked to these diseases, including TGFB1, PIK3CA, and IL-6, suggesting they may serve as therapeutic targets and biomarkers for COPD.

##### 4.1.1.3 Trachea, Bronchus, and Lung (TBL) Cancers

Trachea, bronchus, and lung (TBL) cancers rank among the top ten global causes of death, classified under section 2, Neoplasms, in the ICD-11 [7], highlighting a misalignment with GSEA categories since TBL cancers and general cancers overlap but are not identical. Thus, we made two categories: “Trachea, Bronchus, and Lung (TBL) Cancers” and “Cancers and others.” The TBL cancers, originating from the epithelial lining of the trachea or bronchi, significantly impact the global cancer burden [38,39], with their pathophysiology involving abnormal cell growth and tumorigenesis. Lung carcinoma, which includes small-cell and non-small cell types, is the leading cause of cancer-related mortality [40], while non-small cell lung carcinoma (NSCLC) is the most common subtype, characterized by slower growth compared to small-cell lung cancer; lung adenocarcinoma, a subtype of NSCLC, often affects non-smokers, although smoking remains a significant risk factor [41].

The ICD-11 also categorizes respiratory system (RS) cancers under Section 12 [7], distinguishing them from neoplasms; RS cancers include malignancies affecting the lungs and trachea, with lung cancer under section 2 and bronchial cancers included in the broader lung cancer category due to their common origin in the airways, leading many to progress to higher lung cancer stages [42]. NSCLC encompasses subtypes such as squamous cell carcinoma, large cell carcinoma, and adenocarcinoma, reaffirming the clinical significance of lung adenocarcinoma as a TBL cancer [12]. Thus, these cancers are also included in the TBL cancer category.

##### 4.1.1.4 Ischaemic Heart Disease (IHD)

Ischemic heart disease (IHD) is the leading global cause of death, primarily due to its disruption of coronary circulation through atherosclerosis, which is characterized by plaque buildup [43,44]. It falls under section 11 of the ICD-11 [7], focusing on Diseases of the Circulatory System, which also includes arterial diseases affecting the body’s arteries. A significant example of vascular disease is coronary artery disease (CAD), defined by the accumulation of plaques in coronary arteries, leading to compromised blood flow and directly linked to IHD [45]. Among the diseases identified by gene set enrichment analysis (GSEA), genes such as CETP, TGFB1, APOE, CDKN2B, and among others (Table 1) have been associated with these conditions. The GSEA disease categories largely overlapped with artery disease, hypertension, cardiovascular and cerebrovascular diseases, and stroke, among others.

Additionally, the ICD-11 categorizes cardiovascular system diseases (CVSD) under section 11 [7], which includes complications related to heart and vascular integrity. CAD is responsible for a significant proportion of cardiovascular disease (CVD) cases, accounting for one-third to one-half of presentations [46]. The classification of vascular diseases in this section addresses disorders that affect the vessels of the circulatory system [43–46]. IHD, resulting predominantly from atherosclerosis, reduces oxygenated blood flow to the myocardium and is recognized by the World Health Organization as a major cause of mortality. CAD arises from this atherosclerotic burden, leading to a mismatch between myocardial oxygen demand and supply due to occlusions in coronary vessels [47]. Cardiovascular disease encompasses disorders affecting the heart and vasculature, while vascular disease encompasses any pathology affecting blood vessels, including arteries and veins [46].

##### 4.1.1.5 Stroke

Stroke, characterized by neurological insufficiency due to disrupted cerebral blood flow, is the second leading cause of death globally [1,2]. It is within the ICD-11 under the Diseases of the Nervous System. We found that stroke is covered by cerebrovascular disease [7]. Stroke can occur either through occlusive thrombi or hemorrhagic events caused by ruptured vessels [46]. Thus, it overlaps with GSEA categories discussed in the section on Ischaemic Heart Disease (IHD). The clinical manifestations of stroke include unilateral paralysis, muscle spasms, dysphagia, and various speech and language impairments [47]. Age serves as a critical non-modifiable risk factor, with stroke probability doubling after the age of 55 [48]. Cerebrovascular disease is a broader classification encompassing ischemic events impacting cerebral perfusion, including strokes and aneurysms [49,50].

##### 4.1.1.6 Alzheimer’s Disease (AD)

Alzheimer’s disease (AD) and other forms of dementia are easily categorized among the leading causes of death according to the WHO and the ICD-11 [2,7]. The GSEA disease categories largely overlap with each other, encompassing AD, dementia, neurodegenerative diseases, frontotemporal disease, and amyloidosis. Both AD and other dementias lead to a decline in cognitive function, which significantly affects daily life activities such as thinking, memory, and reasoning. This decline results from neurodegenerative changes in specific regions of the brain [51,52].

The manifestations of dementia can be cognitive, physiological, behavioral, or related to sleep and physical changes [53]. Alzheimer’s disease, one of the most common forms of dementia, affects 10-30% of individuals over the age of 65 and typically progresses over a span of 8-10 years. The disease primarily targets the cerebral cortex and hippocampus [54], disrupting neural networks associated with episodic memory. Both non-pharmacological and pharmacological interventions are aimed at alleviating the suffering associated with Alzheimer’s disease [53]. In general, dementia signifies a clinically observable decline in cognitive function due to brain damage resulting from injury or neurodegenerative disease [55].

##### 4.1.1.7 Diabetes Mellitus (DM)

Diabetes mellitus (DM) was another disease category easily recognized among the leading causes of death according to the WHO and the ICD-11 [2,7]. DM is recognized as the seventh leading cause of death worldwide, according to the World Health Organization. It is classified under section 5 of the ICD-11 [7], which encompasses Endocrine, Nutritional, or Metabolic Diseases. DM primarily consists of two types: type I and type II diabetes. Type I diabetes is usually diagnosed in childhood and occurs due to a complete lack of insulin production. This type requires daily insulin injections, in addition to gene therapy and proper nutritional management [56]. Over time, diabetes mellitus can lead to severe damage and dysfunction of various organs, potentially resulting in organ failure [57].

Hyperglycemia, or elevated blood glucose levels, is classified in the ICD-11 under section 21 [7], which includes symptoms that are not categorized elsewhere. A previous review specifically focuses on severe hyperglycemia, defined as blood glucose levels of 125 mg/dL or higher, with fasting levels reaching 180 mg/dL or more two hours after eating [58]. Although Glucose Metabolism Disease (GMD) is not included in the ICD-11, it refers to conditions where blood glucose regulation is impaired and is closely related to hyperglycemia [59]. Type II diabetes (T2D) is a metabolic disorder characterized by insufficient insulin secretion and insulin resistance in target tissues [60]. Similarly, Carbohydrate Metabolism Disorder (CMD) is not listed in the ICD-11 but affects carbohydrate metabolism and is associated with both diabetes and glycogen storage disorders [61]. Thus, we included those categories in this study.

The GSEA disease categories were insulin-signaling related diseases, including type I and 2 DM, carbohydrate metabolic disorders, and hyperglycemia, among others. The insulin-signaling pathway uses the PI3 kinase-AKT-FOXO signaling, which was identified as a GSEA biological category. Importantly, dysfunction of the insulin signaling pathway causes diabetes, while reducing the pathway functions extends life spans and multiple stress resistance in the model systems [62–65].

##### 4.1.1.8 Kidney Disease, Colonic Disease (CD), and Uterine Disease (UD)

Kidney disease is the eighth leading global cause of death and is classified under Section 16 of the ICD-11 [7], which pertains to Diseases of the Genitourinary System. While kidney disease demonstrated statistically significant results in GSEA, it did not meet our threshold for false discovery rate (FDR). The CD and UD are not classified as kidney diseases; however, there are interesting interactions between them and kidney disease. Thus, we included CD and UD in this category. CD, often referred to as large intestine disease, is listed in section 13 under Diseases of the Digestive System. The inflammatory nature of CD increases the risk of chronic kidney disease, justifying its classification alongside kidney issues [66]. Uterine disease (UD), also found in section 16, includes complications related to the uterus. There is a connection between UD and kidney disease, as kidney issues can disrupt hormonal regulation and lead to abnormal uterine bleeding [67].

##### 4.1.1.9 Cancers and others: General Cancers, Diseases related to Stress Response and Repair, and Progeria

While disease of cellular proliferation (DCP) is not included in the ICD-11, abnormalities in cell growth can lead to various pathologies, particularly cancer [7,68]. Organ system (OR) cancers are closely linked with carcinoma classifications; carcinomas are tumors originating from epithelial cells that can invade locally and metastasize [69,70]. An example is squamous cell carcinoma in the trachea [71]. Key genetic pathways associated with the previously mentioned cancer classifications include TP63, TP53, EGFR, TERT, TNF, and CDKN2A. These genes underscore the shared molecular mechanisms that underlie these malignancies.

The category of autosomal genetic diseases related to age includes a wide range of conditions that impact aging and can be causes of death. This category is particularly enriched with diseases associated with health problems of multiple stress resistance, including organ-specific cancers (like prostate cancer and reproductive organ cancers), diseases with cancer outcomes such as Xeroderma pigmentosum, which is characterized by defects in the DNA repair system It also includes accelerated aging syndromes (such as Cockayne syndrome and progeria) [72], which typically belong to the category of rare diseases.

Importantly, this category indicates a level of resistance to various forms of stress and stimuli (multiple stress resistance), including DNA repair processes, metabolic functions, and cell death [73]. These factors are closely linked to the decisions surrounding cellular survival and death in response to stress.

### 4.2 Methodological advantages and limitations

We utilize GSEA and ORA methods to create a categorization schema that visually represents the varying strengths of these correlations. To better align WHO’s medical-epidemiological categories with genetic ontological categories, we have reclassified the leading causes of death according to their coding, ICD-11. As a result, the WHO’s categories are now more closely aligned with genetic pathway categorizations.

There are limitations primarily arising from the differences between the two categories in the WHO’s leading causes of death and in genetic ontology. One common limitation is the overlap in terminology that is not identical. For example, the mechanisms of ischemic heart disease overlap with stroke within the circulatory system, affecting both the heart and the brain. In another example, trachea, bronchus, and lung (TBL) cancers overlap with general cancer categories; the former specifically impact the respiratory system, while the latter can affect any body system. Thus, we precisely followed ICD-11 to assign the categories from the genetic ontology analysis when possible. For overlapping categories not aligned with the WHO’s categories, we have made the category of Cancers and others.

Another limitation is from the ORA and GSEA methods as discussed previously [5,10,73]. Both analyze gene interactions but use different methods. ORA identifies statistically overrepresented gene sets, highlighting specific biological pathways, while GSEA examines ranked gene lists to find broader associations. By combining both approaches, we gained a more detailed understanding of disease-related gene interactions, revealing distinct pathways and overarching themes. This integrative strategy enhances insights into gene expression dynamics across various diseases. We used the two methods that successfully complemented limitations and identified essential ontological pathways. Relevant to this study, key pathways involved in these categories include PIP3-AKT-FOXO and mTOR, as well as the immune system, metabolism, and autophagy [10]. These pathways largely overlap with mechanisms that confer resistance to various forms of stress [73] and other interventions [74], contributing to healthy aging and increased longevity.

## 5. Conclusions and Perspectives

This study explores the relationship between age-related genetic factors and the primary contributors to global mortality. Our results indicate that age-related genes play a role in all categories of non-communicable diseases, as identified by the leading causes of death, including cancer. The genetic pathway categorizations reveal shared pathways associated with the causes of death, particularly affecting the cardiovascular system, brain, lungs, and the body as a whole. From these results, two important implications arise: (1) Alterations in a single genetic mechanism could impact multiple leading causes of death, making interventions targeting this mechanism potentially effective against these causes; (2) The organ systems affected could serve as targets for clinical diagnosis and interventions aimed at reducing major causes of death and promoting longer lifespans for individuals.

Another important finding is that the genetic contributions go to a wide variety of specific mechanisms as well as a limited number of shared mechanisms. The shared mechanisms overlap with those for exceptional longevity. Despite the prediction of the implausibility of “radical life extension” [75], we predict, based on our observations, that genetic and functional modifying interventions should work to extend lifespans, in the form of genetic, pharmacological, and holistic approaches. For example, extended exposure to endocrine changes confers multiple stress resistance in the cultured primary cells in the long-lived dwarf mice [63,76,77]. Demographic perspectives suggest that the longest-lived populations in specific countries benefit from a wide variety of social, economic, health, cultural, and political factors [78,79].

In conclusion, aging emerges as a pivotal risk factor for a wide array of non-communicable diseases, highlighting its critical influence on health. The decline in immune system function, particularly the reduced responsiveness of B and T lymphocytes, significantly contributes to increased disease susceptibility as individuals age. Shared genes, such as TNF, AKT1, IL6, CDK2A, APOE, and TP53, play vital roles in regulating interconnected inflammatory, metabolic, and growth signaling pathways, reinforcing the concept of a systems-biology approach to understanding aging. Our research indicates that these age-related genes act as pleiotropic hubs, impacting multiple age-related diseases and their underlying pathogenic processes. This interconnectedness underscores the complexity of aging and its health implications, suggesting a need for a holistic view in addressing age-related health challenges.

## Data Availability

All data produced in the present work are contained in the manuscript

